# “Safety and efficacy of pharmacotherapy used for the management of COVID 19: A systematic review and meta-analysis of randomized control trials”

**DOI:** 10.1101/2020.10.02.20206045

**Authors:** Sujit Kumar Sah, Krishna Undela, Sharad Chand, Madhan Ramesh, R Subramanian, Oliver Joel Gona, Sophia M. George, UP Nandakumar, Santosh Aryal, P Niharika, C.S. Shastry

**Affiliations:** Department of Pharmacy Practice, JSS College of pharmacy, JSS Academy of Higher Education and Research, SS Nagar, Mysuru-570015, Karnataka, India; Department of Pharmacy Practice, National Institute of Pharmaceutical Education and Research, Guwahati, Assam, India; Department of Pharmacy Practice, NGSM Institute of Pharmaceutical Sciences, Nitte (Deemed to be University), Paneer, Deralakatte, Mangaluru-575018, Karnataka, India; Department of Pharmacy Practice, JSS College of pharmacy, JSS Academy of Higher Education and Research, SS Nagar, Mysuru-570015; Department of Rheumatology & Immunology, JSS Medical College & Hospital, JSS Academy of Higher Education and Research, SS Nagar, Mysuru-570015, Karnataka, India; Department of Pharmacy Practice, NGSM Institute of Pharmaceutical Sciences, NitteDeralakatte, Mangaluru-575018, Karnataka, India; The Leprosy Mission Nepal, Sunsari and Morang, Nepal; Department of Pharmacy Practice TVM College of Pharmacy, Ballari-583101, Karnataka, India

## Abstract

**Background:** COVID-19 is a novel coronavirus, which is highly contagious and a threat to human health, spreading across nearly 235 countries, affecting 33.8 million and causing 1.01 million fatalities as of 22 September 2020. Researchers have invested tremendous efforts to develop vaccines or effective drug therapy but have not yet been fruitful. Hence, we planned to conduct this systematic review and meta-analysis to supplement the readers with comprehensive data and credible information on the safety and efficacyof essential pharmacotherapy during the pharmacological management of COVID-19.

**Methods:** Theprotocol will be designed based on the updated PRISMA-P 2015 guidelines. An elaborate search of electronic databases such as PubMed/Medline, Web of Science, Scopus, The Cochrane Library, ClinicalTrials.gov, Google Scholar, Medrxiv and other potential databases for articles published during January 2020 to 10 October 2020 is planned to be conducted. Following this,randomized control trials published in English language that assess the safety and efficacy of pharmacotherapy versusplacebo or standard of care or usual care will be evaluated for inclusion. The primary outcomes will include time to clinical recovery and the probability for the negative conversion of COVID-19. Secondary outcomes will quantifythe proportion of patients relieved of symptoms, the all-cause mortality, morbidity, detection of viral RNA, time needed to achieve a negative viral load, ordinal scale changes, ventilatorand oxygen requirements,length of hospital stayand the incidence of adverse and serious adverse events.RevMan V.5.3 computer software packages will be utilised to conduct an accurate statistical analysis of the study. Thebinary random-effects model will be used at a 95 % confidence interval to estimate the weighted effect size ofdichotomous data and continuous data studies. The results of statistical analysis will be considered statistically significant whena p-value <0.05 is attained.

**Results:** Selected studies will be used to evaluate the safety and efficacy of pharmacotherapy used during the management of the novel COVID-19.

**Conclusion:** This study will be a qualitative and quantitative pool of comprehensive evidence regarding the safety and efficacy of pharmacotherapy on COVID-19.

**PROSPERO registration:** CRD42020205433

## Introduction

On 11 March 2020, WHO coronavirus 2019 was declared as a global pandemic and it began to gain worldwide attention. The worsening circumstances required all the domains of health care to proactively engage in the development of immediate and effective strategies to tackle the further spread of the coronavirus. The increase in attention given to the virus was due to itshigh contagious potential accompanied by its enormous threat to human health and the global economy.(1) In December 2019, a new human coronavirus in the Hubei province of Wuhan, China, capable of causing human respiratory disease, severe pneumonia and deathwas first recorded. Soon, COVID-19 spread very rapidly beyond Wuhan to most of the nations resulting in complete locked downsto avoid remission of the virus.(2,3) In February 2020, WHO and the International Committee on Virus Taxonomy named the causative agent as ‘Coronavirus 2019’ and ‘Severe Acute Respiratory Syndrome Coronavirus-2 (SARS-CoV-2)’ respectively.(4,5) Until the 02^nd^ of October 2020, COVID-19 has spread to about 235 countries, affecting nearly 33.8 million people and is responsible for a worldwide mortality of 1.01million.(6)

SARS-CoV-2 is the largest single-stranded, strongly diversified, enveloped, positive-sense RNA virus that is typically found in bats and birds. Transmission to humans occur from these sources often causing acute respiratory distress syndrome (ARDS), which can lead to fatalities.(7,8) The projected mortality rate is an approximate ∼3.8%, which is lower than MERS-CoV (37.1%) or SARS-CoV (10%), but more than ten times the rate of other infections.(9,10) The most frequently observed clinical symptoms in COVID-19 patients are fever, dry cough, dyspnoea, body pain and headache while sore throat, chest pain, diarrhoea, nausea, vomiting, haemoptysis, and conjunctival congestion are less commonly observed. Majority of the confirmed COVID-19 patients are asymptomatic but the potential to transmit the virus continues to be high.(11,12)

RT-PCR and rapid antigen tests by collecting saliva swab tests (91% sensitivity) and nasopharyngeal swab test (98% sensitivity) weredone to detect the presence of the COVID-19 infection.(13)

To provide adequate evidence of the safety and effectiveness of pharmacotherapy, a vast number of clinical trials were quickly registered on ClinicalTrails.gov. Until 2^nd^ October 2020, approximately 3,494 studies related to COVID-19 and 1,976 interventional (clinical trials) studies have been registered in ClinicalTrails.gov. Out of 1,976 interventional trials, 134 had been completed, 74 were terminated, suspended or withdrawn and 21 are in the early phase I trial. 113 studies are currently in the phase I of trials, 270 are in phase II, 163 in phase III and 33 are presently in phase IV.(14) COVID-19 is particularly challenging due to the lack of well-estabalished evidence concerning the safety and effectiveness of therapies and treatment protocolsfor COVID-19. A large number of potential drug candidates for the treatment of COVID-19 such as antiretrovirals: remdesivir, lopinavir/ritonavir, antimalarial: hydroxychloroquine, chloroquine, immunosuppressive agents, angiotensin-converting enzyme 2 (ACE2) inhibitors, non-steroidal anti-inflammatory agents (NSAIDs) and conventional plasma therapy have emerged for use as adjuvants. However, unfortunately, these drug candidates are becoming controversial over time due to various safety and efficacy concerns.(15,16) This may be due to the current lack of a large sample size and the short duration over which studies were conducted. Therefore, we aimed to conduct this systematic review and meta-analysis and expect it to be a credible source ofcomprehensive information on the safety and efficacyof pharmacotherapy during the management of COVID-19.

## Methods

### Protocol registration

Protocol for this study was designed based on the preferred reporting items for systematic review and meta-analysis protocols (PRISMA-P) 2015 statements (17) and has been registered at PROSPERO (CRD42020205433).

### Data sources

We will conduct a comprehensive search of electronic databases such as PubMed/Medline (https://www.ncbi.nlm.nih.gov/pubmed/), Web of Science (https://clarivate.com/web-of-science-core-collection/), Scopus (https://www.scopus.com/), The Cochrane Library (http://www.cochranelibrary.com/), ClinicalTrials.gov (https://www.clinicaltrials.gov/), Google Scholar (https://scholar.google.com/), Medrxiv (https://www.medrxiv.org/) and other potential databases. Articles published during January 2020 to 10 October 2020 will be included for analysis. The study is planned to be completed by 30^th^ October, 2020. The references of key articles will be identified through hand searches. The human studies (randomized control trials) published in English language that evaluated the safety and efficacy of pharmacotherapy versus placebo or standard of care or usual care will be considered for review as well.

### Search strategies

Key words or relevant texts that will be used to conduct a rigorous comprehensive literature search are pharmacotherapy, COVID-19 and study designs including drug therapy, antiviral agents, remdesivir, GS-573, lopinavir-ritonavir drug combination, oseltamivir, dolutegravir, indinavir, arbidol, antimalarials, hydroxychloroquine and chloroquine were reviewed for analysis. Immunosupressive agents, glucocorticoids, dexamethasone, antirheumatic agents, tocilizumab, sarilumab, biological products, antibodies, monoclonal, anti-bacterial agents, azithromycin, nafamostat, camostat, nitazoxanide, famotidine, angiotensin-converting enzyme inhibitors, non-steroidal anti-inflammatory agents were some other key words that were used while probing the databases. Serotherapy, immunization, vaccines, COVID-19, 2019 novel coronavirus infection, coronavirus disease 2019, 2019-nCov infection, SARS-CoV-2, severe acute respiratory syndrome coronavirus-2, SARS2, 2019, randomized control trial, clinical trials and RCTs were other terms that were used to make sure that no eligible articles were excluded from review.

The following search strategies will be adopted to search PubMed databases presented below:

#1. Search “drug therapy”[MeSH Terms]

#2. Search ((((((((((((((((((((((((((((antiviral agents) OR remdesivir) OR lopinavir-ritonavir drug combination) OR oseltamivir) OR indinavir) OR arbidol) OR antimalarials) OR hydroxychloroquine) OR chloroquine) OR immunosupressive agents) OR glucocorticoids) OR dexamethasone) OR antirheumatic agents) OR tocilizumab) OR sarilumab) OR biological products) OR antibodies) OR monoclonal) OR anti-bacterial agents) OR azithromycin) OR nafamostat) OR camostat) OR nitazoxanide) OR famotidine) OR angiotensin-converting enzyme inhibitors) OR non-steroidal anti-inflammatory agents) OR serotherapy) OR immunization) OR vaccines

#3. Search #1 OR #2

#4. Search “COVID-19” [Text Word]

#5. Search (((((2019 novel coronavirus infection) OR coronavirus disease 2019) OR 2019-nCov infection) OR severe acute respiratory syndrome coronavirus-2) OR SARS2) OR 2019-nCoV

#6. Search #4 OR #5

#7. ((Randomized control trial) OR clinical trials) OR RCTs

#8. Search #3 AND #6 AND #7

In order to conduct the search withinother scientific databases, this search strategy will be adopted and therelevant key terms will be amended as necessary.

### Eligibility Criteria

Eligibility for this study will be evaluated based onthe PICOS (populations, interventions, comparison,outcomes, and study designs) format.

### Study population

➢ Patients who have a positive COVID-19 diagnostic report using the RT-PCR or serological methods
➢ Patients of any age, gender and race
➢ Subjects at any clinical stage of illness (mild to serious conditions) are eligible for inclusion
➢ Patients with or without other comorbidities

### Intervention

➢ Drug therapy or vaccines of any dose and combinations

### Comparison

- Placebo or standard care or usual care

### Outcome measures

Both primary and secondary outcomes will be given importance. The primary outcomes will include the time to clinical recovery and the probabilityof a negative conversion of COVID-19. The secondary outcomes include a computation of the proportion of patients who experienced symptomatic relief, a measurement of all-cause mortality, morbidity, detection of viral RNA, time taken to achieve negative viral load, frequency of progression to respiratory illness, quantified body temperatures and time taken to recover from cough. Aditionally, impact on laboratory and radiological parameters, ordinal scale changes, ventilation and oxygen requirements, ICU admission rate, duration of hospitalization, time for discharge, rate of development of severe infections or complications, incidence of adverse outcomes (common and rare) and serious adverse events were also among the other listed secondary outcomes that would be measured by the study. The results of these consequences will be statistically describedas mean, median, percentile and number of days (such as on the 7^th^, 14^th^ and 28^th^ day).

### Study Designs

➢ Randomized control trials

## Exclusion criteria

- Wrongly diagnosed or an unconfirmed diagnosis of COVID-19
- Patients using ayurvedic and homeopathic agents or any other form of conventional treatment
- Literatures such as non-randomized control trials, observational studies, retrospective studies, commentaries, notes and abstracts will be excluded.
- Paper reports with a total sample siz eof less than 10 recruits.
- Records with missing information or those that were unable to gather information

### Study selection

The records will be retrieved from the above mentioned databases by using the above described search strategies. The extracted data will be transferred to Mendeley for the removal of duplicates. After the elimnation of duplicates, titles and abstracts will be screened and reviewed independently by three authors to check for the satisfaction of the outlined eligibility criteria. Each review author will be blinded towards any information about journals and authors of the paper. Any disagreement during the selection process while checking for the eligibility of studies will be resolved through discussion with a fourth author.

### Data extraction

Three reviewers will independently extract qualitative datafrom the selected studies that have mentioned the first author of the study, publication year, country, study setting, study design with the blinding status specified, patient’s demographics details (age and gender), total sample size, sample in intervention and control group, loss of follow-ups, details on interventions (such as drug name, dose, duration of therapy), comparators (placebo or standard care or usual care), duration of follow-ups, outcomes measured (both primary and secondary) and risk of bias. In order to summarize the findings, changes that have occurred from baseline till the end of follow-up,the difference in changes within groups and between groups, significance level at 95% confidence interval and the status of findings that werefavourable for the intervention group will be quantifed and mentioned. The data will be extracted into Microsoft Excel and transferred into suitably designed data collection forms in Microsoft word. Following which, the results will be tabulated as tables and expressed as figures. After data extraction by independent review authors is completed, it will further be exchanged for critical analysis of the relevant information so that blinding will be eliminated. Any disagreements during the extraction of data will be resolved through constructive dialogue and discussions with the fourth author.

### Assessments of reporting risk of bias

Three review authors will assess the risk of bias independently using the Cochrane Risk of Bias (RoB). The risk of bias will be categorised as ‘low’, ‘unclear’ and ‘high’ based on essential domains including randomization sequence generation, concealment of allocation, blinding of participants and personnel, blinding of outcomes assessment, incomplete outcome data, selective reporting, and other bias. The decision on the risk of bias will be transferred to computer-based RevMan V.5.3. in order to generate an image of risk of bias graph and risk of bias summary. Any conflicts on judgment will be resolved through conversations among the authors of the review.

### Statistical analysis

In this review, the scope for meta-analysis is limited due to the wide range of outcomes that are evaluated. However, if at least two studies indicate homogeneity with similar interventions and outcomes, a meta-analysis will be performed. RevMan V.5.3 computer software packages will be used to perform all statistical analysis. The dichotomous data obtained will be used to quantify the occurring events while mean with standard deviations will be provided to represent continuous data. The binary random-effects model will be used at a 95 % confidence interval to estimate the weighted effect size of studies which will be summarised using risk ratios. Both the Chi-squared test and I^2^ testing will be done to estimate the heterogeneity between studies. An I^2^ value>50% will be described as the presence of moderate to high heterogeneity. All statistical results will be considered statistically significant when the p-value is measured to be less than 0.05.

### Sub-group analysis

A subgroup analysis will be carried out in the presence of relevant data. The analysis carried out might include subgroups of studies conducted country-wise, subject characteristics and the overall adverse effects of drugs.

### Sensitivity analysis

After a subgroup analysis is performed, if a substantial heterogeneity is noticed in the included trials then additional tests will be carried out to further investigate the source of heterogeneity for this systematic review. If more than ten studies will be available then a funnel plot will be drawn to analyse for the risk of reporting bias.

### Addressal of missing data

If any relevant data is missing or incomplete, review authors will engage themselves to actively search for the original sources or contact authors through mail to obtain any missing information. If it is not possible for missing datato be retrieved, then the study will be excluded from the analysis.

### Ethics and dissemination

In this review, previously published data will be used for the synthesis of data. Therefore, provision for ethical approval and informed consent will not be required. This study is scheduled to be written and will be published or presented in peer review journals and conferences.

### Disclosures of funding

This study was not funded or supported by any organization or industry which may have influenced this study at any stage of review.

## Discussion

Most of the potential drug therapies rapidly went under trial because of the observed beneficial effects during the occurrence of previous epidemics such as MERS-CoV and SARS-CoV. Some drugs have shown beneficial effects during in-vitro preclinical trails.(18) However, safety and efficacy issues such adverse effects, a nil reduction of viral load, drug-drug interactions for majority of the drugs remain a cause of controversy and they are highly likely to be ineffective. Hence, it is required to pay more attention to these facets in order to establish the safety and efficacy of pharmacotherapy used for the treatment of COVID-19. The main objective of this study is to provide comprehensive and accurate evidence on the security and therapeutic efficacy of available pharmacological measures. The authors of this review firmly believe that the most promising therapeutic strategies to control and cure COVID-19 can be concluded from the findings of this review. This will enableus to offer guidance for the future clinical management of the confirmed COVID-19 cases.

**Figure 1:**
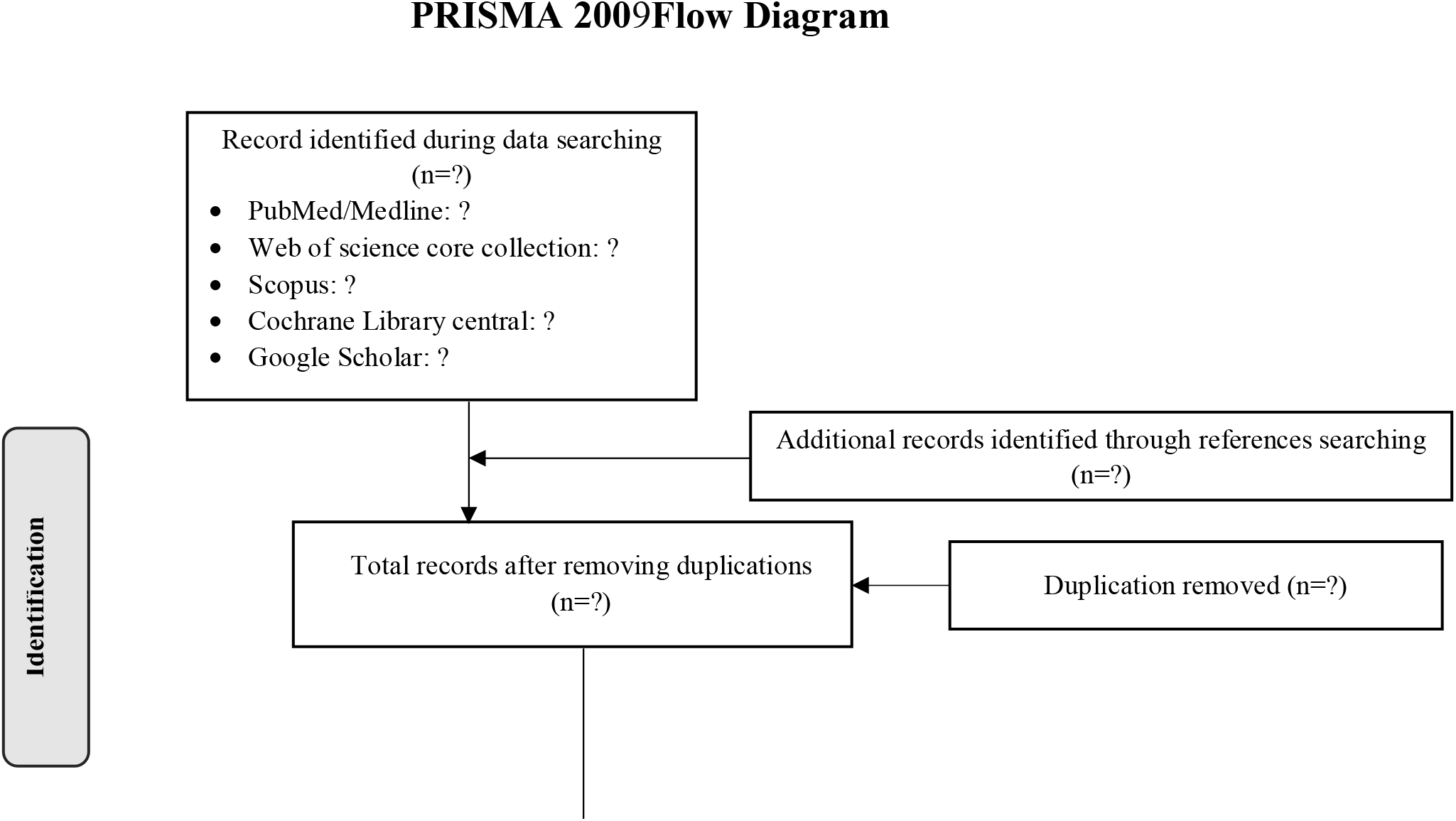
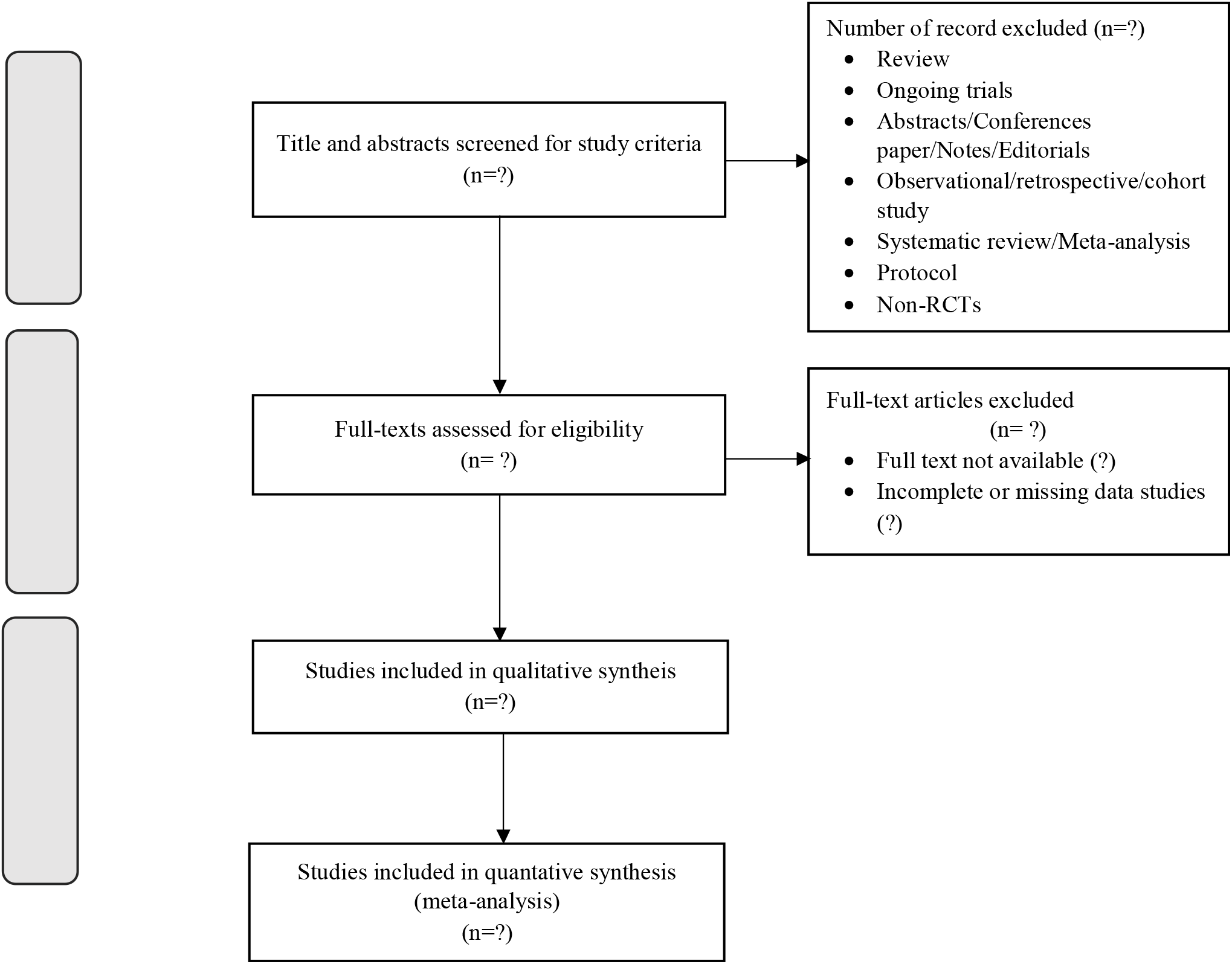
Flowchart of Study Selection Process (PRISMA)

## Data Availability

The additional data will be provided on request

